# Forecasting glucose from CGM and sparse meal logs with a residual-gated multimodal transformer

**DOI:** 10.64898/2026.06.01.26354646

**Authors:** Jung Lee, Suheng Yao, Lin Tang, Xiaowei Yu, Michael C. Cheney, Honghuang Lin, Xuezhou Zhang, Debarghya Mukherjee, Maura E. Walker, Nicole L. Spartano, Huimin Cheng

**Author notes:** Corresponding author: Huimin Cheng, August 30, 2026.

## Abstract

Forecasting glucose from continuous glucose monitoring in free-living settings is challenging because trajectories depend on endogenous dynamics and sparsely recorded meals. We developed MealRes-Gate, a multimodal transformer that produces a complete CGM-based forecast and then adds a gated, meal-informed residual correction. The correction is controlled by combined self-reported and CGM-derived evidence of meal presence, scaled per forecast horizon, allowing for dietary context to improve predictions without requiring meal records. We evaluated MealRes-Gate in 1,752 adults without diabetes from the Framingham Heart Study. Participants wore Dexcom G6 Pro sensors and completed paired ASA24 dietary recalls. MealRes-Gate achieved the lowest mean absolute error across forecast horizons, meal-state strata, and glucose ranges compared with long short-term memory, transformer, and GluFormer model architectures. It also improved prediction of postprandial peak glucose, time-to-peak, positive incremental area under the curve, and time in the 70–140 mg/dL range. Participant-level cross-validation confirmed generalization to unseen participants.

## 1 Introduction

Glucose dysregulation is central to many metabolic diseases, including diabetes and its vascular complications [1]. Because diabetes develops progressively, the prediagnostic period provides an important opportunity for prevention. In the Diabetes Prevention Program, intensive lifestyle intervention reduced diabetes incidence by 58% among highrisk adults [2]. These findings underscore the importance of characterizing glucose dynamics before overt diabetes develops, particularly among non-diabetic individuals.

Continuous glucose monitoring (CGM) captures glucose under free-living conditions [3, 4, 5, 6, 7, 8], but remains largely retrospective. Forecasting glucose 30 minutes to 2 hours ahead could provide early warning of impending excursions and support timely behavioral decisions [9, 10, 11, 12, 13, 14, 15, 16]. For example, after a meal, a forecast may identify continued glucose rise before the peak is visible on the CGM trace. Accurate forecasting in free-living individuals is nevertheless difficult because free-living glucose reflects the interaction of endogenous physiological dynamics with incompletely observed behavioral exposures. Dense CGM measurements make recent glucose history highly predictive at short horizons, whereas longer-horizon forecasts increasingly depend on exogenous perturbations, particularly meals [5, 15, 17, 18, 19].

Meals are a major source of acute glucose excursions, with effects that can persist for several hours [5, 17, 19, 20, 21]. Unlike CGM, which is recorded automatically, dietary intake is typically self-reported and therefore sparse, incomplete, and inconsistent across participants [22, 23, 24, 25, 26]. This mismatch between densely observed CGM trajectories and sparsely observed meal records creates a key challenge for multimodal forecasting: meal information may be highly informative when available, but models cannot assume its availability [3, 4, 5, 15].

Existing approaches do not fully address this challenge. Prior work on glucose forecasting has explored classical autoregressive and related time-series methods [12, 27, 28, 29], early neural network and conventional machine learning approaches [30, 31], deep re-current neural networks [32, 33, 34, 35], and more recent transformer-based architectures [36, 37, 38, 39, 40, 41]. Models incorporating dietary information have also demonstrated that meal information can improve glucose prediction, but most meal-augmented approaches have been developed in diabetes cohorts where meal logging is relatively complete due to its tie to insulin management [32, 35, 42, 43, 44]. Conventional multimodal fusion approaches that concatenate modalities, impute missing observations, or otherwise assume comparable modality availability [45, 46, 47, 48, 49, 50], are poorly suited to free-living glucose forecasting, where CGM is continuously observed but dietary information is systematically sparse. What is needed is not simply another multimodal forecasting architecture, but a framework that explicitly considers the unequal reliability of its inputs. CGM should provide a robust forecast on its own, while dietary information should refine the forecast selectively with additional evidence.

To address this problem, we developed MealRes-Gate, a residual-gated multimodal transformer organized around an explicit hierarchy of modality reliability. The model first forms a CGM-based forecast from short-term and long-term glucose dynamics, then applies dietary information as an optional, horizon-specific correction to that forecast. A combination of self-reported meal record and a CGM-derived estimate of meal presence activates the correction when the glucose trace itself carries a meal signal, so the correction can respond to physiological evidence when a self-report is missing. This design matches what free-living data provide, which is a dense passive glucose measurement paired with sparse behavioral report. Dietary information improves the forecast when it is informative, and CGM remains the primary forecasting signal when no meal is reported.

We evaluated MealRes-Gate using data from 1,752 adults without diabetes in the Framingham Heart Study (FHS). Participants wore Dexcom G6 Pro CGM devices for up to 10 days under free-living conditions and completed paired Automated Self-Administered 24-Hour (ASA24) Dietary Assessment Tool recalls [51, 52]. The model predicted 24 future 5-minute glucose measurements spanning four forecast horizons: 0–30, 30–60, 60–90, and 90–120 minutes. We compared MealRes-Gate with long short-term memory (LSTM) network [32, 35], transformer [53, 36, 38], and a GluFormer model retrained in our data using the architecture of the recently proposed CGM foundation model [41]. Evaluation included both a chronological within-participant design, representing forecasting for an individual with previously observed data, and five-fold participant-level cross-validation (CV), representing deployment to individuals not encountered during training. In addition to overall forecasting accuracy, we assessed performance in postprandial and non-postprandial windows, low-, mid-, and high-glucose states, and across demographic and metabolic subgroups.

MealRes-Gate achieved the lowest forecasting error across all evaluated horizons and physiological subgroups, with its advantage becoming more pronounced as the prediction horizon increased and in the more difficult postprandial and extreme-glucose states. The improvements also translated to more accurate prediction of postprandial peak glucose, time-to-peak, positive incremental area under the curve (pAUC), and time in the 70–140 mg/dL range. MealRes-Gate also maintained its performance advantage when evaluated on held-out participants, indicating that the learned forecasting patterns generalized beyond participants represented during training. This work establishes a methodological foundation for future systems that seek to move CGM from retrospective monitoring toward proactive, personalized metabolic guidance before overt diabetes develops.

## 2 Results

The study included 1,752 FHS participants with normoglycemia or prediabetes who wore Dexcom G6 Pro sensors for up to 10 days and completed ASA24 dietary recalls on two weekdays and one weekend day (Figure 1a). This protocol produced an asymmetric multimodal dataset where CGM recorded glucose every 5 minutes, whereas dietary intake was reported intermittently (Figure 1b). We aligned both modalities to consecutive 30-minute windows. Each window contained a 60-dimensional CGM feature vector of raw glucose values and summary features of shortand long-term glucose dynamics, and a 20-dimension dietary feature vector of nutrient measures and meal-context variables (Figure 1c). Full feature and model specifications are given in the Methods (Sections 4.3 and 4.6). From these aligned input sequences, the model predicts six 5-minute glucose values within each of four successive forecast windows, 0–30, 30–60, 60–90, and 90–120 minutes, for a total of 24 future values.

**Figure 1:**
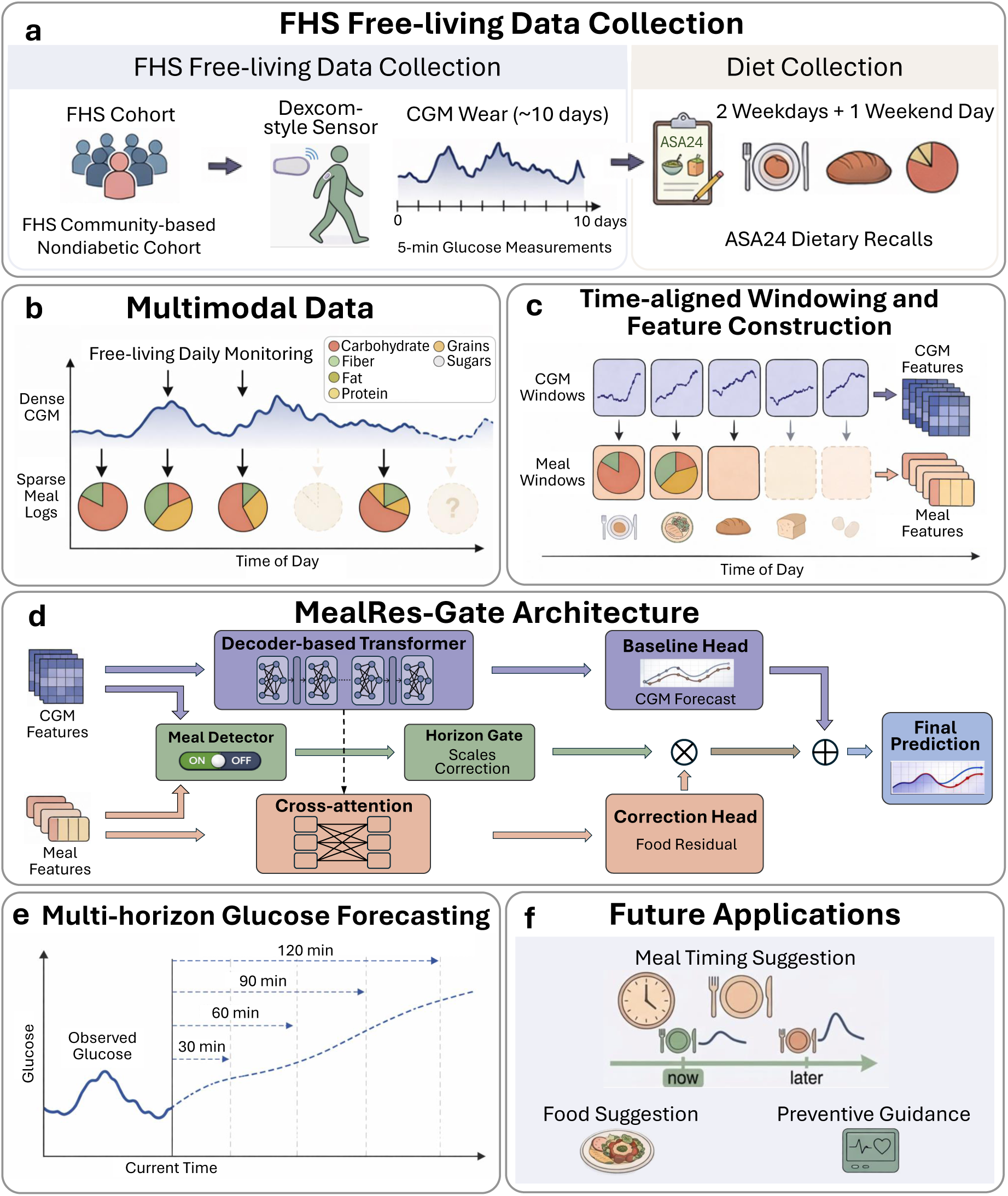
Free-living multimodal data collection and MealRes-Gate workflow. **a,** In the Framingham Heart Study (FHS) free-living setting, continuous glucose monitoring (CGM) was collected continuously whereas dietary intake was recorded intermittently. **b,** Glucose trajectories are dense and meal logs are sparse with heterogeneous macronutrient composition. **c,** CGM and meal records were aligned into 30minute windows and converted into separate CGM and meal feature representations. **d,** MealRes-Gate uses a transformer CGM backbone to generate a baseline forecast and an auxiliary meal branch with cross-attention, meal detection, and horizon-specific gating to produce a residual meal correction. **e,** The model predicts 24 future 5-minute glucose measurements spanning four forecast horizons. **f,** Forecasts may support meal guidance and timing recommendations.

The computation of the predictions proceeds in five steps (Figure 1d). (1) A transformer backbone encodes the input-window CGM sequence and a baseline head generates 24 future values. (2) A separate meal pathway applies cross-attention between the aligned dietary sequence and the CGM representation, producing a 24-value residual correction. (3) A CGM-based meal-detection module uses engineered CGM features to estimate the probability of a concurrent meal, *P* (meal). The area under the receiver operating characteristic curve (ROC-AUC) of this module is 0.705, showing that CGM dynamics contain useful information for detecting meal presence (Methods Section 4.5; Supplementary Figure S5). The estimated probability is then combined with the self-reported meal indicator, taking the larger of the two values, to form an effective meal-presence signal. (4) This signal and a horizon-specific gate, driven by meal timing and size, scale the residual correction, allowing the influence of dietary information to vary with both evidence of a meal and forecast horizon. (5) The scaled correction is added to the CGM-only forecast to produce the final prediction. During training, prediction errors are minimized using a zone-weighted smooth *L*_1_ loss that assigns greater importance to errors in clinically important and underrepresented low and high glucose ranges, preventing the more abundant mid-range observations from dominating model learning. The resulting trajectory spans the 30-, 60-, 90-, and 120-minute horizons (Figure 1e). Figure 1f illustrates potential downstream uses, including meal-selection and meal-timing guidance. These applications were not directly evaluated in the present study.

### 2.1 Study population and free-living data

We analyzed CGM data from the FHS, a multigenerational, community-based cohort with extensive phenotypic and clinical characterization. The present study included participants from the Third Generation, New Offspring, and Omni 2 cohorts who attended their fourth examination cycle between September 2022 and July 2025 (*n* = 2,718). The design for these FHS cohorts has been described previously [54]. Participant inclusion and exclusion criteria for the current analysis are summarized in Figure 9 and detailed in Methods Section 4.1.

Under this protocol, 2,147 participants wore a Dexcom G6 Pro CGM device on the abdomen or the upper arm for up to 10 days under free-living conditions, with interstitial glucose recorded every five minutes (FDA 510(k) K191833). During the same monitoring window, dietary intake was assessed using the ASA24, a validated web-based dietary assessment system developed by the National Cancer Institute [51, 52]. Participants were asked to complete two weekday recalls and one weekend recall, and reported meal events were mapped to the nearest 5-minute CGM interval to align dietary information with the glucose time series. After applying device-related, data-quality, and clinical exclusion criteria (Figure 9), 1,752 participants were included in the final analysis. We used two evaluation protocols, a within-participant chronological split and a five-fold participant-level CV, described in Sections 2.2 and 2.7, respectively, and illustrated in Supplementary Figure S1.

### 2.2 Multi-horizon glucose forecasting

We first evaluated MealRes-Gate in the setting where future glucose is predicted for participants with historical records already available. For each participant, we applied a chronological split of the longitudinal data into training (60%), validation (20%), and test (20%) segments, so that model fitting and hyperparameter tuning used only earlier observations and all reported results were obtained on a held-out future segment, with test-set forecasts using only preceding observed CGM and meal history as input, as would be available at deployment. Forecasting was formulated as a sliding-window task such that at each eligible anchor time *t*, the model consumed the preceding 6 hours of CGM and meal data and predicted glucose over the subsequent four horizon windows (0–30, 30–60, 60–90, and 90–120 minutes). We evaluated performance using window-averaged MAE, the mean absolute error across the six 5-minute glucose measurements within each 30-minute horizon window, averaged across all evaluated horizon windows (Supplementary Figure S3).

To evaluate performance under physiologically distinct conditions, we stratified the 447,504 forecast windows in the held-out test set by meal context and glucose range (Supplementary Figure S2). Postprandial and non-postprandial classifications were restricted to valid-diet days, defined by sex-specific bounds on total reported daily energy intake (Methods Section 4.4.1). This restriction made the absence of a logged meal more plausibly interpretable as a non-meal period. A forecast window was classified as postprandial if a caloric meal (kcal *>* 0) was recorded within the 2 hours preceding that window or within the window itself, else, it was classified as non-postprandial. Under this definition, 14,518 windows (3.2%) were postprandial and 18,224 (4.1%) were non-postprandial. The remaining 414,762 windows (92.7%) were from ineligible diet-days and were reported as a robustness stratum rather than as a meal-defined group.

Each 30-minute forecast window was also assigned to one mutually exclusive glucose stratum using its six observed target values. A window was classified as low glucose if any value was below 70 mg/dL, high glucose if any value exceeded 140 mg/dL and none was below 70 mg/dL, and mid glucose (70–140 mg/dL) otherwise. Low glucose was given priority when both low and high values occurred within the same window. This classification yielded 2,256 low-glucose windows (0.5%), 336,626 mid-glucose windows (75.2%), and 108,622 high-glucose windows (24.3%). We used 140 mg/dL rather than the conventional diabetes-focused threshold of 180 mg/dL to capture elevated glucose excursions in this cohort of adults without diabetes.

Across all six panels of Figure 2, forecasting error increased with prediction horizon for every method, as expected. MealRes-Gate achieved the lowest MAE in every stratum and at every horizon, although the margin over the best competing baseline was small and depended somewhat on the horizon. At the 0–30 min horizon, MealRes-Gate’s advantage over the best competing baseline was under 1 mg/dL in every stratum. It grew at the 90–120 min horizon to 1.21 mg/dL in postprandial windows, 1.22 mg/dL in non-postprandial windows, 2.05 mg/dL in low glucose windows, and 1.88 mg/dL in high glucose windows. The wider gap at the longer horizons is where the gated meal correction and the engineered CGM features that encode historical glucose trends add information that raw CGM history alone does not capture.

**Figure 2:**
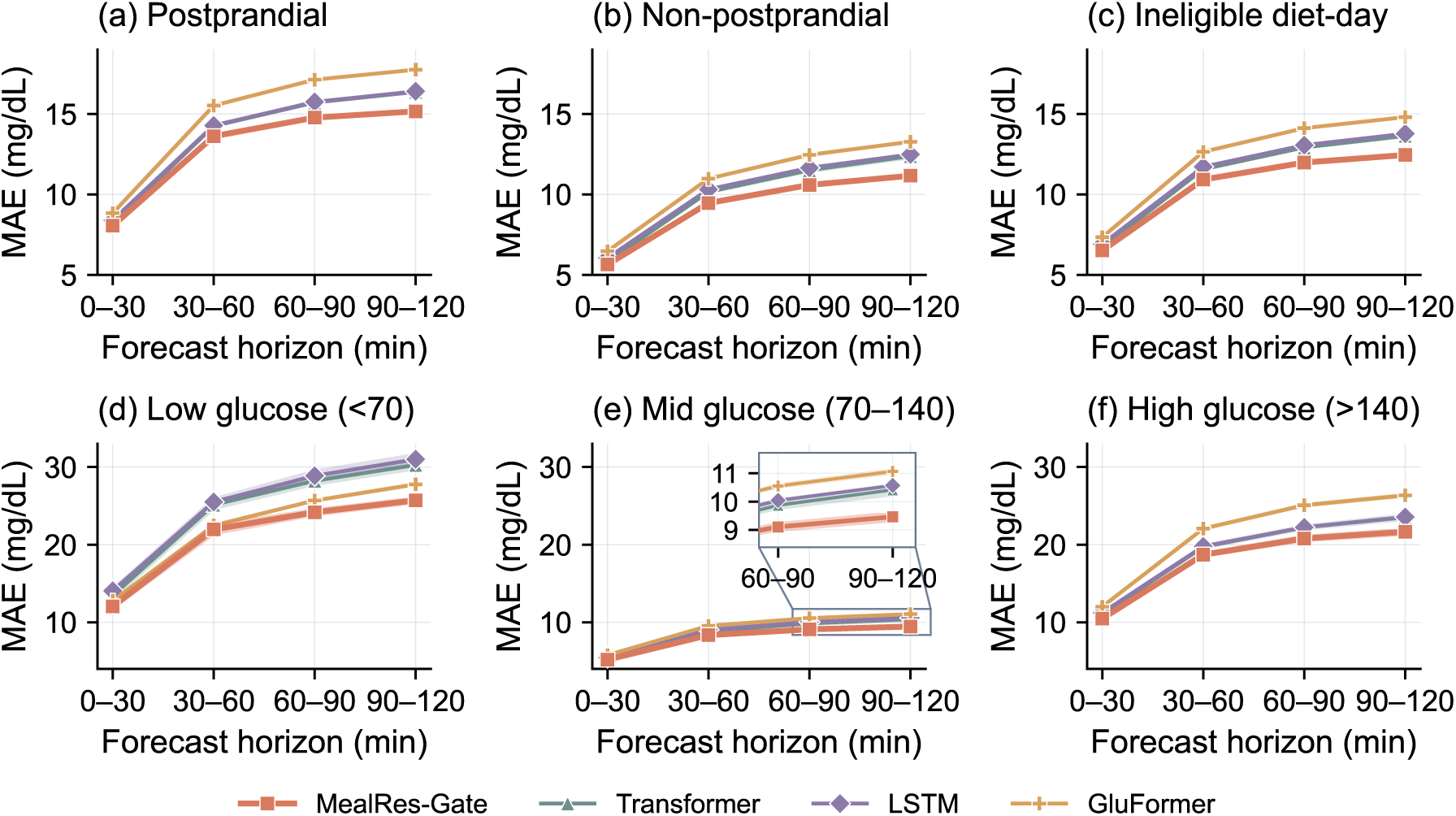
Forecasting performance across meal-context and glucose strata. Window-averaged MAE at the 0–30, 30–60, 60–90, and 90–120-minute forecast windows for MealRes-Gate and the competing methods. Panels **a** and **b** are restricted to validdiet days. **a,** Postprandial (PP) windows, those with a caloric meal in the 2 hours ending at that horizon. **b,** Non-postprandial (non-PP) windows, those with no such meal. **c,** Ineligible diet-day windows, horizons on days outside the valid-diet-day filter and therefore, ineligible for inclusion as either postprandial or non-postprandial. Panels d-f are stratified based on the observed target glucose values in each forecast window. **d,** Low glucose, any value below 70 mg/dL. **e,** Mid glucose, all values within 70– 140 mg/dL. **f,** High glucose, any value above 140 mg/dL and none below 70 mg/dL. Lines show the mean across five random seeds and shaded bands indicate ±1 standard deviation across seeds.

Postprandial windows were consistently more difficult to forecast than non-postprandial windows (Figure 2a,b). For MealRes-Gate, MAE was 8.05 mg/dL in postprandial windows and 5.64 mg/dL in non-postprandial windows at 0–30 minutes, a difference of 2.41 mg/dL. At 90–120 minutes, the corresponding errors were 15.16 and 11.16 mg/dL, increasing the gap to 4.00 mg/dL. This pattern is consistent with the additional uncertainty introduced by variation in meal timing, quantity, composition, and individual postprandial response. Importantly, MealRes-Gate also remained the best-performing method in non-postprandial windows, indicating that its benefit was not restricted to periods with an explicitly recorded meal.

The largest absolute errors, and the clearest separation between methods, occurred in lowand high-glucose windows (Figure 2d,f). At 90–120 minutes, MAE ranged from approximately 26 to 31 mg/dL in low-glucose windows and from 22 to 26 mg/dL in high-glucose windows across the evaluated methods. The difficulty of these strata likely reflects both their rapid, nonlinear dynamics and the influence of unmeasured behavioral and physiological factors. Low-glucose windows were also rare in this cohort, limiting the number of examples available for model training. Despite these challenges, MealRes-Gate maintained the lowest mean error in both extreme-glucose strata.

We further tested whether MealRes-Gate significantly outperformed the comparison models within each forecast horizon and across meal-context and glucose strata (Supplementary Table S4). Statistical testing procedures are described in Section 4.10.1. Most comparisons met the prespecified significance criterion, providing statistical support for the lower forecasting error observed with MealRes-Gate.

### 2.3 Ablation study

We next conducted ablation studies to evaluate how model components contributed to forecasting performance (Figure 3). AB1 replaced the composite training loss with standard mean squared error (MSE), removing both the glucose zone and horizon weights and the velocity-consistency component (Method 4.7). AB2 removed the CGM-derived meal probability, *P* (meal), leaving meal presence to be determined only by the selfreported meal indicator while retaining the horizon-specific gate (Method 4.5). AB3 removed all explicit dietary features (Method 4.3.1). AB4 removed the engineered CGM feature set, leaving only raw CGM input (Method 4.3.2). Each heatmap cell reports the change in MAE relative to the full model, calculated as ablation minus full-model MAE. Therefore, positive values (red color) indicate worse performance after removing the component, suggesting that the component contributes to forecasting performance.

**Figure 3:**
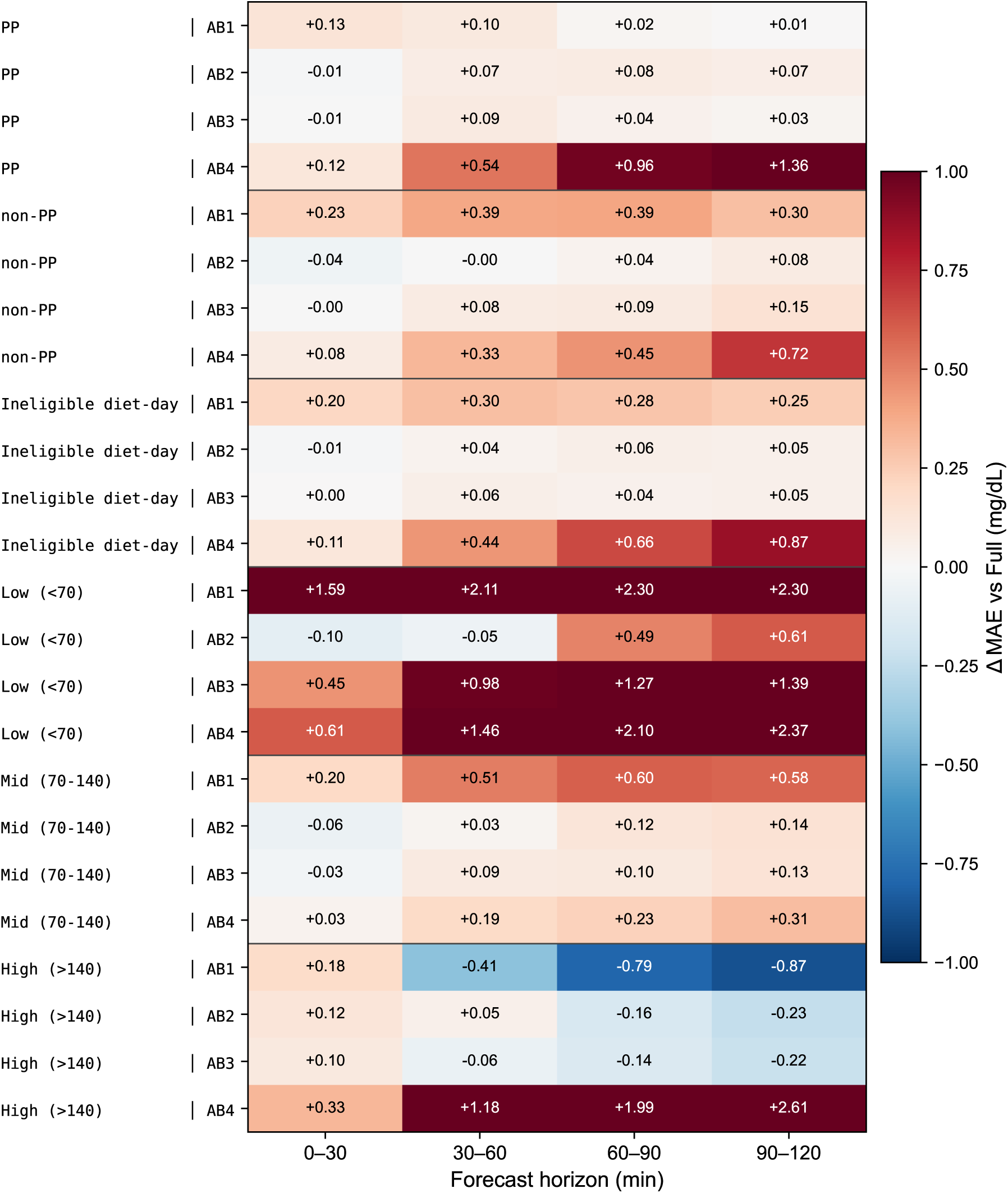
Component ablation across meal-state and glucose subgroups. Change in window-averaged MAE relative to the full MealRes-Gate model (ablation minus full), averaged over eligible windows within each subgroup and horizon and then across five training seeds. Meal-state (PP, non-PP, Ineligible diet-day) and glucose subgroups (Low, Mid, High) follow the definitions in Figure 2. Red denotes higher MAE than the full model (worse performance), and blue denotes lower MAE. The color scale is saturated at ±1.0 mg/dL, while each cell reports the exact value.

#### Zone-weighted loss primarily improves low-glucose forecasting

Replacing the composite loss with standard MSE (AB1) increased low-glucose MAE by 1.59, 2.11, 2.30, and 2.30 mg/dL across the 0–30, 30–60, 60–90, and 90–120-minute windows, respectively. Because the composite loss includes a glucose-zone-specific weighting component, the improvement in the low-glucose stratum is most likely attributable to this zone weighting. These increases were consistent across all five training seeds and were substantially larger than the AB1 effects in the other subgroups. More modest increases appeared in non-postprandial, ineligbile diet-day, and mid-glucose windows, whereas postprandial MAE changed by only 0.01–0.13 mg/dL. In high-glucose windows, standard MSE produced lower mean MAE at the three longer horizons (−0.41, −0.79, and −0.87 mg/dL), but the direction of these differences was not consistent across seeds (Supplementary Table S6).

#### The CGM-derived meal probability improves longer-horizon forecasting

The effect of removing *P* (meal) (AB2) increased with forecast horizon. Across the postprandial, non-postprandial, and ineligible diet-day subgroups, mean ΔMAE increased from +0.01 mg/dL over the two shorter-horizon windows (0–30 and 30–60 minutes) to +0.06 mg/dL over the two longer-horizon windows (60–90 and 90–120 minutes).

This horizon-dependent benefit was most pronounced in low-glucose windows, where mean ΔMAE changed from −0.07 mg/dL over the two shorter-horizon windows to +0.55 mg/dL over the two longer-horizon windows. Thus, the CGM-derived meal probability provides a modest, horizon-dependent refinement in that becomes more useful when forecasting beyond 60 minutes.

#### Explicit dietary features contribute primarily beyond the 30-minute horizon

Removing all explicit dietary features (AB3) produced essentially no average change at 0–30 across the meal-state subgroups, with a mean ΔMAE of 0.00 mg/dL. In contrast, the mean ΔMAE across 30–60, 60–90 and 90–120 was +0.07 mg/dL, and removing these features increased MAE in 15 of the 18 displayed subgroup cells at these horizons. These results indicate that dietary context provides a modest horizon-dependent correction beyond the meal-presence signal, with its contribution becoming more apparent as the influence of recent meals unfolds over time.

#### Engineered CGM features were the dominant source of predictive gain

Removing the engineered CGM representation (AB4) produced the largest and most consistent degradation across strata, with effects that increased sharply with horizon. In high-glucose windows, MAE increased by 0.33, 1.18, 1.99, and 2.61 mg/dL across the four forecast windows. In low-glucose windows, the corresponding increases were 0.61, 1.46, 2.10, and 2.37 mg/dL. Postprandial MAE increased by 0.12, 0.54, 0.96, and 1.36 mg/dL.

### 2.4 Performance across participant subgroups

To assess robustness across participant subgroups, we computed each participant’s average MAE across all windows and stratified it by demographic and clinical characteristics, including sex, age, body mass index (BMI), prediabetes status, glucose coefficient of variation, and percentage of CGM values above 140 mg/dL (Figure 4). Participant counts per subgroup are reported in Supplementary Table S7.

**Figure 4:**
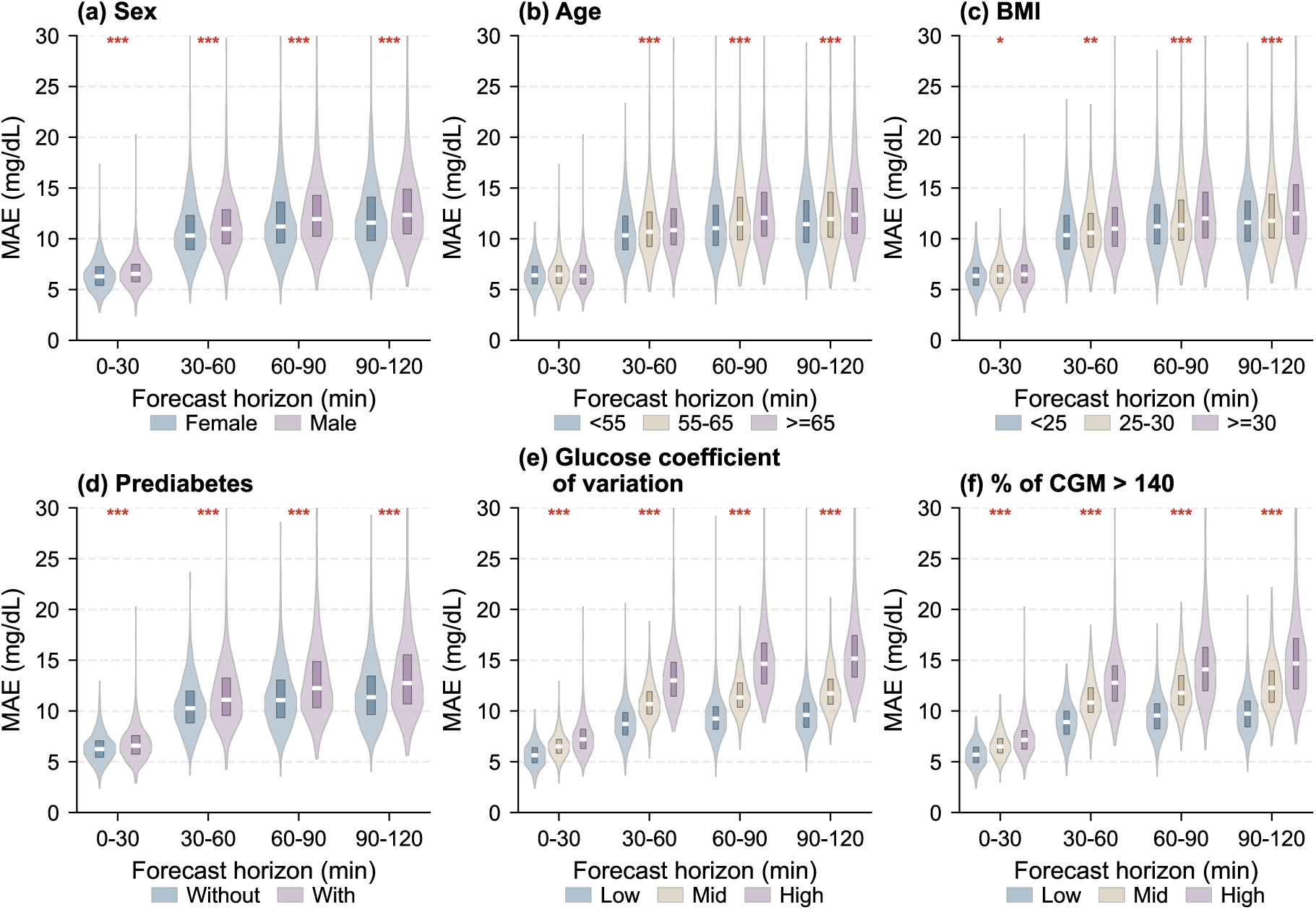
Subgroup analysis of forecasting performance across demographic and clinical strata. Distribution of participant-level mean window-averaged MAE for MealRes-Gate across four successive 30-minute forecast windows, stratified by (a) sex, (b) age, (c) BMI, (d) prediabetes status, (e) tertile of glucose coefficient of variation, and (f) tertile of the percentage of CGM *>* 140 mg/dL. Participants were classified as without prediabetes based on HbA1c *<* 5.7 and fasting venous plasma glucose *<* 100 mg/dL [55]. Significance markers (* *p <* 0.05, ** *p <* 0.01, *** *p <* 0.001) denote Benjamini–Hochberg-adjusted *p*-values from the 24 primary subgroup tests (six subgroup dimensions across four forecast windows). Non-significant comparisons are not annotated. See Methods Section 4.10 for the statistical analysis and Supplementary Table S7 for subgroup sample sizes. Results are from a single training run.

Across all strata, MAE increased monotonically with prediction horizon, consistent with the greater uncertainty associated with longer-range glucose forecasting. Differences across demographic factors were comparatively modest in magnitude. The sex-, age-, and BMI-stratified panels showed broadly overlapping error distributions, with only small differences in median per-participant MAE of 0.92 mg/dL or less at the 90–120 min horizon (small relative to the overall increase in error across prediction horizons). This pattern suggests that the model is reasonably robust across these participant characteristics and maintains broadly consistent performance across sex, age, and BMI subgroups.

In contrast, larger differences emerged when participants were stratified by metabolic status. Individuals with prediabetes and higher glucose coefficient of variation showed systematically higher MAE at longer horizons. At the 90–120 min horizon, median per-participant MAE increased from 11.36 mg/dL among participants without prediabetes to 12.74 mg/dL among those with prediabetes, and from 9.62 mg/dL in the low glucose coefficient of variation subgroup to 15.16 mg/dL in the high glucose coefficient of variation subgroup, a gap of 5.54 mg/dL that exceeds the differences observed across any demographic subgroup. Participants who spent a greater proportion of time above 140 mg/dL showed a similar pattern.

These findings suggest that participants with greater glycemic instability and more time spent at elevated glucose levels exhibit more complex glucose dynamics that make future trajectories inherently harder to forecast, and that remaining prediction difficulty is driven more by metabolic heterogeneity than by demographic characteristics alone.

### 2.5 Model explanations and feature importance

We used permutation importance to characterize the CGM and dietary features that contributed most strongly to prediction at the 0–30 and 90–120-minute horizons (Figure 5). For each feature, values were randomly permuted across forecast windows while all other inputs and targets were held fixed [56]. Importance was defined as the percentage increase in MAE relative to the non-permuted model. Each permutation was repeated ten times and averaged. Under this definition, an importance of 100% indicates that permuting the feature doubled the baseline MAE.

CGM feature importance, evaluated across all windows, shifted markedly with forecasting horizon. At the 0–30 min horizon (Figure 5a), prediction was driven mainly by 24-hour mean glucose (+31.6%), followed by the most recent 5-minute glucose rate of change (+20.4%), current glucose at window *t* (+14.5%), and glucose at t−5 minutes (+14.4%). This indicates that 0–30 min horizon forecasting depends on both the individual’s baseline glycemic level over the preceding 24 hours and the current trajectory with its recent rate of change.

**Figure 5:**
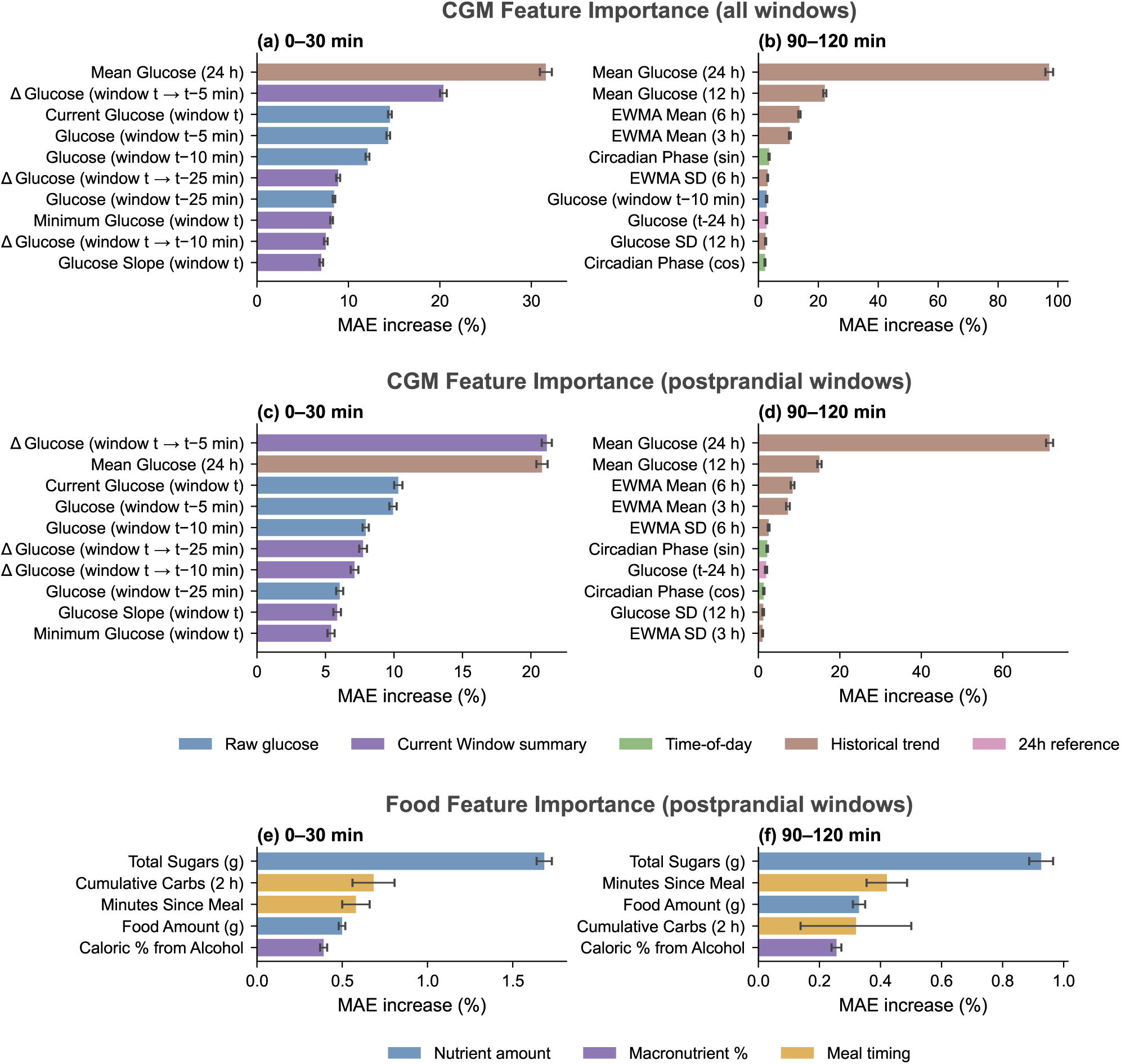
Feature importance of CGM and food features at short and long forecast horizons. Permutation importance of the top 10 CGM features and top 5 food features, measured as the percentage increase in MAE after feature permutation. Panels **a,c,e** show the 0–30 min horizon and panels **b,d,f** show the 90–120 min horizon. **a,b,** CGM features across all test windows. **c,d,** CGM features across postprandial windows. **e,f,** Food features across postprandial windows. Bars show the mean over ten permutation draws, with error bars indicating ±1 standard deviation across those draws. Results shown are from a single training run. Equivalent panels for the remaining four seeds are provided in Supplementary Figures S6–S9.

At the 90–120 min horizon (Figure 5b), the ranking changed substantially. The 24hour mean glucose remained important but became clearly dominant (+97.1%, indicating that permuting it nearly doubles the baseline MAE at this horizon), followed by broader historical summaries such as 12-hour mean glucose (+22.1%) and exponentially weighted moving averages with 6-hour and 3-hour half-lives (+13.7% and +10.5% respectively), as well as the circadian phase encoding. At long horizons, the model thus relies less on immediate local fluctuations and instead anchors its prediction to the average glucose over the past day, adjusting based on accumulated trend and circadian context.

Restricting the CGM permutation analysis to postprandial windows (Figure 5c–d) shifts the short-horizon ranking. At the 0–30 min horizon the most recent 5-min glucose rate of change becomes the single most important feature (+21.2%), narrowly ahead of the 24-hour mean glucose (+20.8%), with the current glucose slope entering the top ranks (+5.9%). At the 90–120 min horizon the 24-hour mean glucose again dominates (+71.5%), as in the all-window analysis. This suggests that immediately after a meal, short-horizon forecasting leans more on recent glucose dynamics, while long-horizon forecasting still anchors to the participant’s average glucose over the preceding day.

These horizon-dependent patterns were consistent across the four additional training seeds (Supplementary Figures S6–S9), with short-horizon rankings consistently emphasizing both recent glucose dynamics and 24-hour mean glucose, whereas long-horizon rankings were dominated by 24-hour mean glucose and other historical summaries.

Food feature importance was evaluated on postprandial windows only, where dietary effects are most pronounced (Figure 5e–f). We restricted the permutation analysis to the nutrient and meal-timing features, excluding the binary meal indicator and the CGM-derived *P* (meal). These two features are involved in the multiplicative gate that controls the food pathway and therefore, permuting them generates input combinations the model never encountered during training (e.g., the gate opens on an empty food presence or closes on a true meal), inflating their apparent importance above what a feature-level interpretation supports. The contribution of the gating signal as a whole is instead quantified via the AB2 structural ablation in Section 2.3.

Among the remaining food features, total sugars (+1.68%), cumulative carbohydrate intake over the preceding 2 hours (+0.68%), and minutes since last meal (+0.58%) were the most influential at the 0–30 min horizon (Figure 5e), reflecting the immediate glycemic impact of sugar and carbohydrate content and the recency of food consumption. Carbohydrate-related features remained important at the 90–120 min horizon (Figure 5f), where total sugars (+0.93%), minutes since last meal (+0.42%), and food amount (+0.33%) were the largest contributors, consistent with the sustained glycemic effect of carbohydrate-rich and larger meals [57]. Although the exact feature ranking varied across training seeds, carbohydrate-related features were consistently represented among the top five food features at both horizons in the four supplementary analyses. These included total sugars, cumulative carbohydrate intake, net carbohydrates, carbohydrate percentage, and refined grains, supporting a consistent carbohydrate-centric dietary signal across training runs.

Other macronutrient composition features (alcohol, protein) contributed smaller but non-negligible signal. Apart from total sugars at the short horizon, these per-feature effects were small in absolute terms, on the order of 1% or below. This is consistent with the ablation result that explicit food features provide a modest refinement rather than a dominant signal and reflects that permutation importance measures the marginal contribution of a single dietary feature while the gating pathway is held fixed.

### 2.6 Forecasting clinically relevant postprandial metrics

We next asked whether improved glucose forecasting translates into more accurate prediction of clinically relevant postprandial summaries. We evaluated four derived outcomes computed cumulatively from the start of the forecast to each horizon (0–30, 0–60, 0–90, 0–120 min): peak glucose, time-to-peak, pAUC, and time-in-range 70–140 mg/dL (Figure 6)[58, 59]. Peak glucose and time-to-peak characterize the magnitude and timing of the postprandial excursion, pAUC measures the integrated glucose excursion above a pre-forecast baseline and is reported in time-normalized form so that values are comparable across cumulative horizons. Time in range of 70–140 mg/dL is the percentage of the forecast period during which glucose remains between 70 and 140 mg/dL. Full metric definitions are given in the Methods (Section 4.9.2).

**Figure 6:**
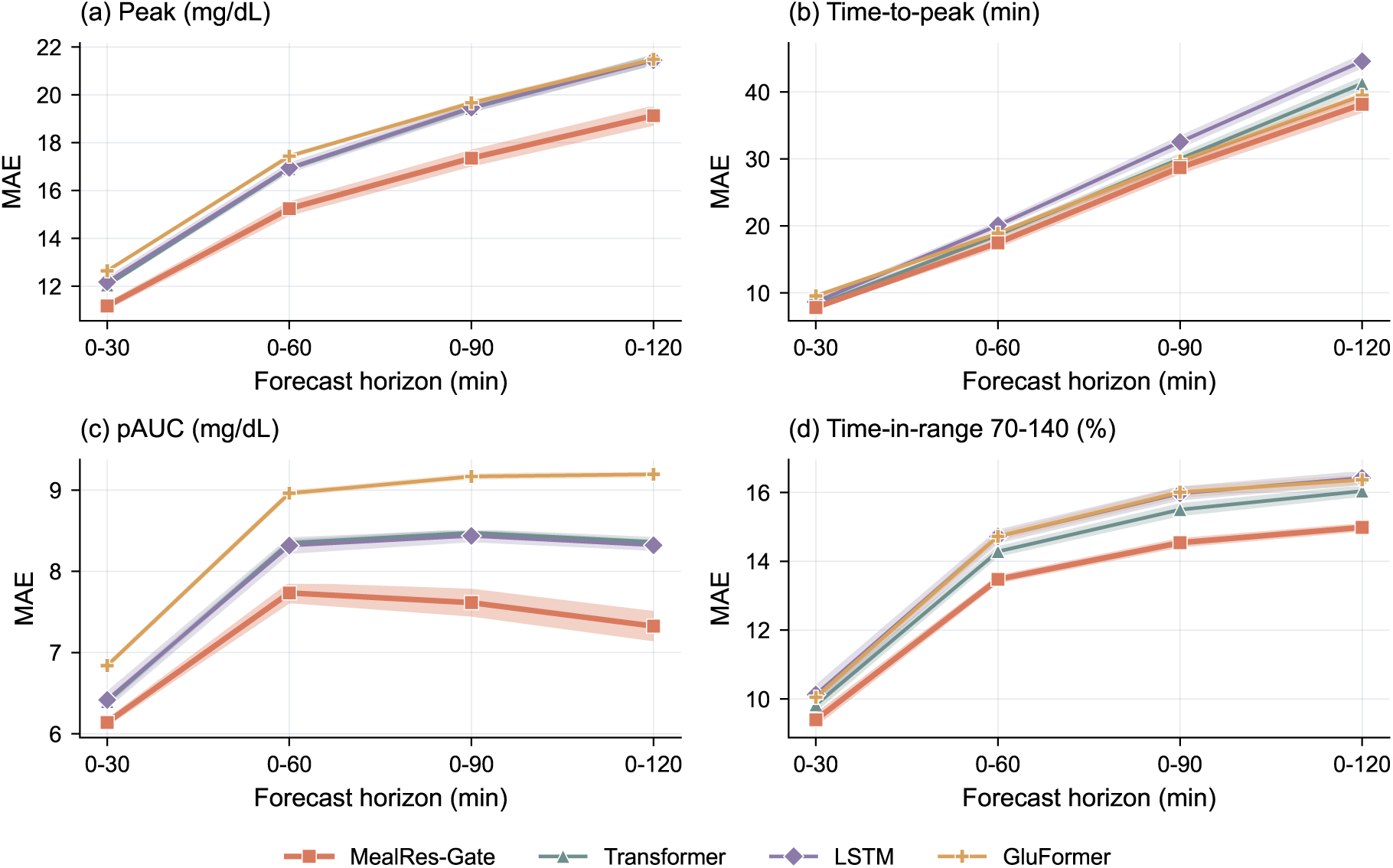
Forecasting accuracy for four derived postprandial clinical out-comes. MAE between outcomes computed from predicted and observed glucose trajectories over cumulative forecast horizons (0–30, 0–60, 0–90, and 0–120 min). **a,** Peak glucose. **b,** Time-to-peak. **c,** positive incremental area under the curve (pAUC). **d,** Time-in-range 70–140 mg/dL. Analyses include only postprandial forecast trajectories. Peak glucose and pAUC are reported in mg/dL, time-to-peak in min, and time-in-range in percentage points. Lines show the mean across five random seeds, and shaded bands indicate ±1 standard deviation across seeds.

For this analysis, we included the 2-hour forecast trajectories in which all four consecutive 30-min windows met the previously defined postprandial criterion. Unlike glucose MAE, which can be evaluated separately within the 0–30, 30–60, 60–90, and 90–120 min windows, these clinical outcomes summarize the trajectory from the forecast start through a given horizon. Calculating the clinical metrics separately within each 30-min window would fragment this response because peak glucose would represent a local rather than overall maximum, time-to-peak would no longer describe the timing of the overall postprandial peak, pAUC would capture only part of the cumulative glucose exposure, and time in range would reflect only a brief portion of the response. We therefore calculated these metrics over cumulative horizons of 0–30, 0–60, 0–90, and 0–120 min.

MealRes-Gate achieved the lowest MAE at every cumulative horizon across all four postprandial metrics (Figure 6). Improvements were most pronounced for peak glucose, pAUC, and time in range and generally increased at longer horizons. In contrast, the four methods achieved broadly comparable time-to-peak performance, although MealRes-Gate retained the lowest MAE. Among the competing methods, performance rankings varied by outcome. GluFormer had the highest MAE for peak glucose and pAUC, whereas LSTM had the highest time-to-peak MAE.

The comparable time-to-peak performance did not imply comparable trajectory accuracy. Figure 7 shows three illustrative forecasts in which MealRes-Gate and GluFormer predicted similar peak times but produced markedly different trajectories. MealRes-Gate more closely followed the observed glucose rise, peak magnitude, and subsequent trajectory, whereas GluFormer showed larger discrepancies in excursion magnitude and shape. Time-to-peak captures only when the maximum occurs and does not characterize the magnitude, cumulative exposure, or overall shape of the postprandial response. These examples therefore support evaluating peak glucose, time to peak, pAUC, and time in range together.

**Figure 7:**
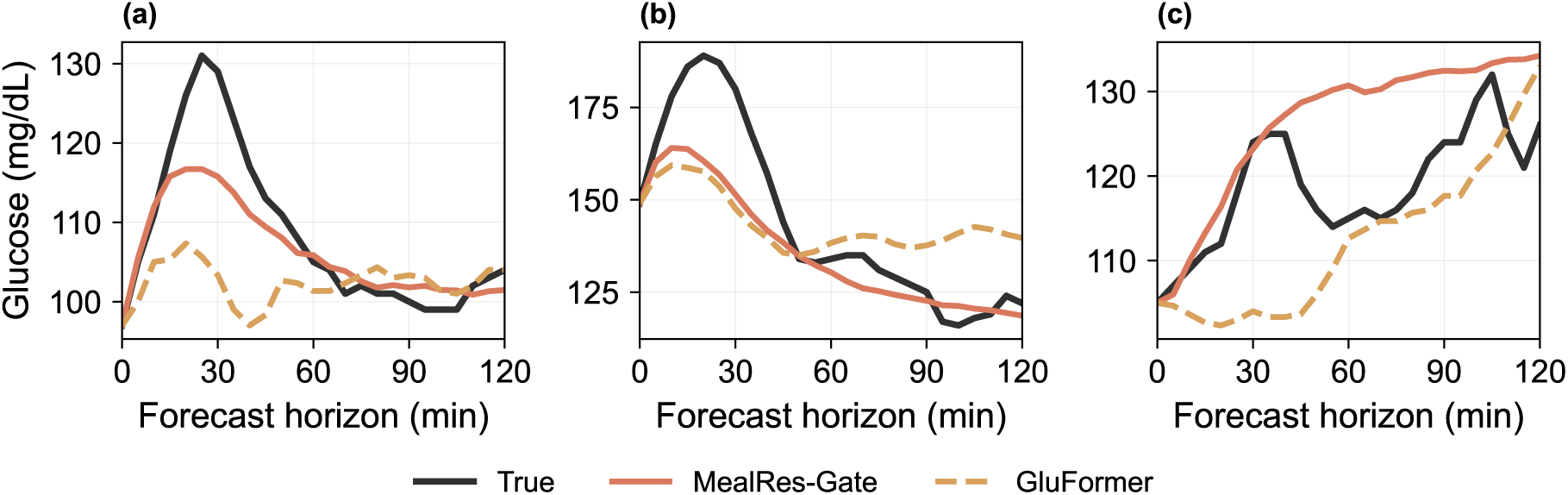
Similar time-to-peak estimates can mask differences in postprandial trajectory accuracy. Three illustrative 2-hour postprandial forecasts in which MealRes-Gate and GluFormer predicted similar times to peak but different trajectory shapes. Black lines show observed glucose, solid red lines show MealRes-Gate predictions, and dashed orange lines show GluFormer predictions.

Error patterns across cumulative horizons also differed by metric. Peak-glucose and time-to-peak MAE increased as the cumulative horizon expanded, reflecting the greater difficulty of predicting the magnitude and timing of a peak over a longer trajectory. For MealRes-Gate, pAUC MAE shows a rise-then-slight-decline pattern, increasing from 6.14 mg/dL at 0–30 min to 7.73 mg/dL at 0–60 min, then decreased slightly to 7.33 mg/dL at 0–120 min. Because time-normalized pAUC represents the average positive glucose excursion per minute, later portions of the trajectory with smaller prediction discrepancies can reduce the cumulative average error. When the additional pAUC discrepancy grows more slowly than the duration of the evaluation window, errors accumulated during the earlier rise and peak are averaged over a longer period, resulting in a lower normalized pAUC MAE. Time-in-range MAE increased most sharply between 0–30 and 0–60 min and more gradually thereafter, as each additional glucose value has progressively less influence on the cumulative percentage at longer horizons.

### 2.7 Generalization to unseen participants

The evaluations in the preceding subsections address a within-participant forecasting setting, where earlier observations are used for training and later observations are used for testing. This design is appropriate for assessing prospective prediction over time but does not address a distinct and practically important question of whether the model can generalize to *new individuals* whose data were never observed during training. In CGM-based digital health applications, this distinction is critical. A model may perform well when it can learn participant-specific baselines and response patterns from historical data, yet degrade substantially when deployed to a previously unseen person with different metabolic dynamics, meal-reporting behavior, and glucose variability. Evaluating generalization to unseen participants is therefore necessary to determine whether the model captures transferable physiological structure rather than relying primarily on participant-specific memorization. To assess generalization, we performed five-fold participant-level CV. In each fold, 80% of participants were used for training, and the remaining 20% were split equally into validation (10%) and test (10%) sets, ensuring that all three sets were fully disjoint at the participant level.

As shown in Figure 8, MealRes-Gate achieves the lowest MAE at every horizon and in every clinical subgroup, indicating that its gains are not merely due to repeated exposure to the same participants. In postprandial windows (Figure 8a), MealRes-Gate reaches 15.49 ± 0.54 mg/dL at the 90–120 min horizon, compared to 16.38–17.95 mg/dL across the competing methods (transformer, LSTM, and GluFormer). A comparable gap persists in non-postprandial windows (Figure 8b), where MealRes-Gate (10.50 ± 0.24 mg/dL at the 90–120 min horizon) outperforms the next-best method, the transformer (11.98 ± 0.19 mg/dL).

**Figure 8:**
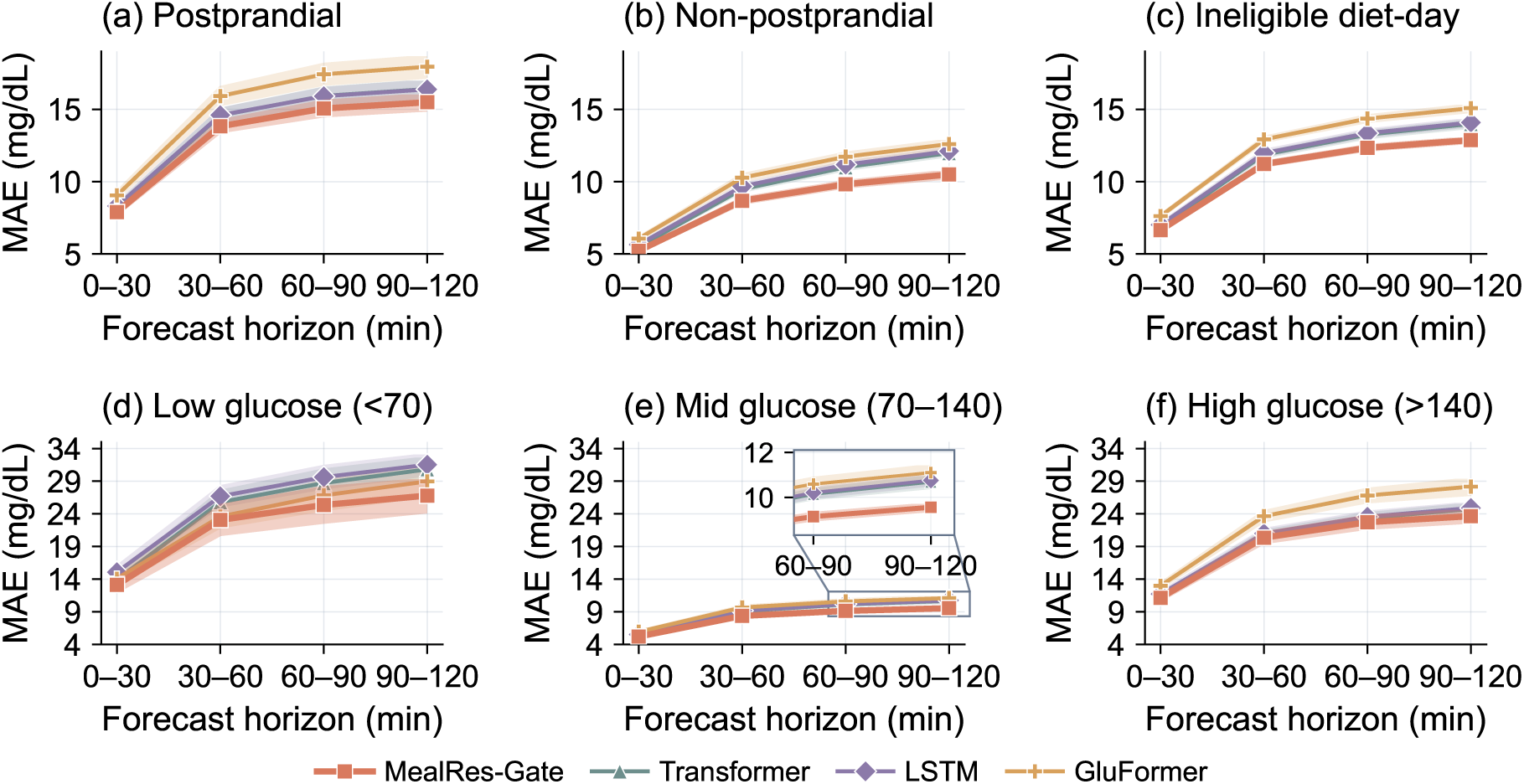
Generalization to unseen participants under five-fold participant-level CV. Window-averaged MAE at each prediction horizon for MealRes-Gate, transformer, LSTM, and GluFormer. Meal-state subgroups (postprandial, non-postprandial, other), and glucose subgroups (low, mid, high) are defined as in Figure 2. Shaded bands indicate ±1 standard deviation across the five folds. Each fold represents a single training run using the same fixed random seed.

**Figure 9:**
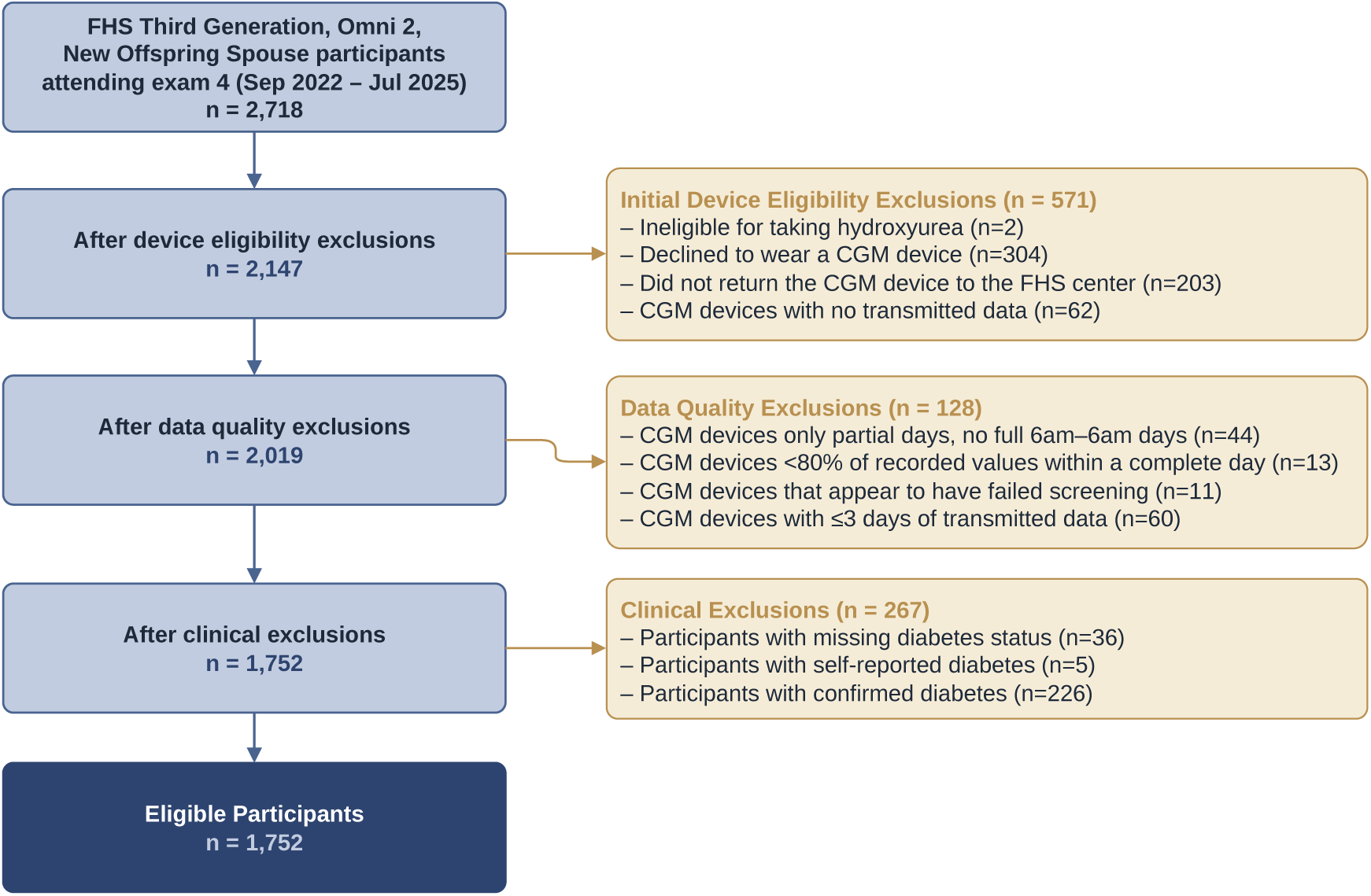
Participant selection flowchart. From an initial 2,718 Framingham Heart Study participants, sequential device-related, data-quality, and clinical exclusions yielded the final analytic cohort of 1,752.

The advantage was also retained in the more difficult extreme-glucose strata. At the 90– 120 min, high-glucose MAE was 23.65 ± 0.93 mg/dL for MealRes-Gate, compared with 24.76–28.17 mg/dL for the competing methods. In low-glucose windows, MealRes-Gate achieved 26.80 ± 2.51 mg/dL, compared with 28.97–31.53 mg/dL. Cross-fold variability was larger in the low-glucose stratum because these events were uncommon, but MealRes-Gate retained the lowest mean error.

We similarly tested whether MealRes-Gate outperformed the comparison models within each forecast horizon and across meal-context and glucose strata under participant-level cross-validation (Supplementary Table S5). Statistical testing procedures are described in Section 4.10.1. Most comparisons met the prespecified significance criterion, providing statistical support that the lower forecasting error of MealRes-Gate extends to participants who were not observed during training.

## 3 Discussion

In this study, we developed MealRes-Gate, a residual-gated multimodal transformer for forecasting free-living glucose trajectories in adults without diabetes and with prediabetes. Across both the within-participant chronological evaluation and participant-level CV, MealRes-Gate achieved the lowest mean absolute error among the evaluated recurrent, transformer, and GluFormer architectures at every forecast horizon and across all reported meal-context and glucose strata. The advantage was modest over the first 30 minutes but increased at longer horizons. Improvements also extended beyond pointwise glucose values to postprandial trajectory summaries, including peak glucose, time-to-peak, pAUC, and time in the 70–140 mg/dL range. Together, these findings indicate that free-living glucose forecasting benefits from an architecture that treats continuously observed CGM as the primary predictive signal and incorporates sparse dietary information only as a constrained refinement.

The gain was not limited to pointwise error. MealRes-Gate also reproduced the shape of the postprandial response more closely, improving peak glucose, cumulative positive glucose exposure, and time in the 70–140 mg/dL range. Even though the differences in time-to-peak were small, our illustrative trajectories show that excursions with nearly the same time-to-peak can differ substantially in peak magnitude and in overall shape. Evaluating these outcomes jointly is therefore important: time-to-peak captures when the maximum occurs, but not the magnitude, duration, or cumulative burden of the response.

Glucose forecast difficulty varied with both forecast horizon and metabolic state. Prediction error increased with forecast horizon for all methods, was consistently higher in postprandial than non-postprandial windows, and was greatest in lowand high-glucose states. At short horizons, glucose continuity and recent rates of change strongly constrain the near-term trajectory. The widening separation between methods at longer horizons therefore suggests that multiscale historical information and contextual refinement become more useful when simple extrapolation from the most recent glucose values is no longer sufficient. At the participant level, performance was comparatively stable across sex, age, and BMI, whereas larger differences were associated with prediabetes, glucose variability, and the proportion of readings above 140 mg/dL. This suggests that forecasting difficulty is driven more by metabolic instability and dynamic glycemic state than by the demographic characteristics.

The feature-importance analyses further revealed that the information supporting glucose prediction changes substantially with forecast horizon. At 0–30 minutes, prediction depended on both the individual’s recent glycemic state and immediate glucose dynamics. In postprandial windows, the 5-minute rate of change became the leading short-horizon predictor, consistent with the immediate trajectory carrying substantial information once a meal-related excursion has begun. At 90–120 minutes, however, prediction shifted away from local fluctuations toward broader historical context: the 24-hour mean glucose became overwhelmingly dominant, followed by the 12-hour mean, multihour exponentially weighted averages, and circadian phase.

Among dietary features, total sugars was the strongest predictor at both forecast horizons, although the ranking of the other features varied. During the first 30 minutes, cumulative carbohydrate intake over the previous 2 hours ranked second, followed by time since the meal and food amount. Shortly after eating, the quantity and type of rapidly absorbable carbohydrate therefore provide the most direct information about the developing glucose response. Between 90 and 120 minutes, time since the meal became the second most important dietary feature, followed by food amount, while cumulative carbohydrate intake became less prominent. This shift suggests that longer-horizon prediction depends increasingly on where an individual lies along the postprandial trajectory, i.e., whether glucose is still rising, near its peak, or returning toward baseline. Food amount may also help indicate how large the response will be and how long it will last.

Despite these findings, our study has some limitations. First, the present study evaluated generalization to previously unseen participants within the FHS rather than transportability across independent cohorts. Participant-level cross-validation ensured that individuals in the test sets were entirely absent from model training, providing evidence that the observed performance was not solely attributable to memorization of participant-specific glucose baselines or daily rhythms. Cross-cohort validation, however, addresses a more stringent and distinct question. Independent populations may differ in demographic and metabolic characteristics, dietary behavior, sensor platforms, monitoring duration, and data-collection protocols, all of which could induce distribution shifts that are not represented by within-cohort validation. Rigorous external evaluation therefore requires independent free-living cohorts containing temporally aligned CGM and detailed dietary measurements. Such datasets remain difficult and costly to collect and were not available for the present study. Future work should evaluate MealRes-Gate across populations, study designs, and CGM platforms and determine when direct transfer is adequate versus when cohort-specific recalibration or model adaptation is required.

Second, dietary intake was assessed using self-reported ASA24 recalls, which may be incomplete and subject to errors in meal timing, portion size, and nutrient composition. Although MealRes-Gate was designed to accommodate sparse meal information, more timely and detailed dietary measurements may further improve forecasting. Future studies could combine self-reported dietary records with image-based dietary assessment to capture meals closer to the time of consumption and provide complementary information about food type and portion size.

Third, the present model considered CGM and dietary information but did not incorporate several other determinants of short-term glucose dynamics. Physical activity, sleep, heart rate, stress, illness, circadian patterns, and medication use may explain part of the remaining prediction error, particularly during periods in which glucose behavior cannot be explained by recent CGM and meals alone. The residual-gating formulation provides a natural mechanism for extending the model to these additional modalities: each source could contribute a separately gated residual component while preserving CGM as the continuously available predictive backbone. Such extensions should, however, be evaluated incrementally. Adding modalities increases measurement burden, missingness, and model complexity, and additional information should not be assumed to improve prediction merely because it is available.

Beyond forecasting accuracy, an important downstream application is the development of anticipatory decision-support systems that help individuals understand how nearterm behaviors may affect their glucose trajectories. For example, a forecasting model could continuously update an individual’s expected glucose trajectory and identify periods in which a planned meal is likely to produce an unfavorable postprandial response. Rather than providing a single generic recommendation, such a system could compare feasible alternatives, for example, changes in meal composition, portion size, timing, or post-meal activity, and present options predicted to produce more favorable glucose profiles. This would shift CGM from a predominantly retrospective monitoring tool toward a prospective system that supports decisions before an undesirable glucose response occurs.

A particularly promising extension is to combine MealRes-Gate with sequential decisionmaking methods such as reinforcement learning. In such a framework, the individual’s recent CGM history and behavioral context could define the state, modifiable behaviors such as meal composition, meal timing, or physical activity could constitute candidate actions, and the reward could reflect multiple objectives, including postprandial glucose excursions, time in a target range, nutritional adequacy, behavioral burden, and adherence. MealRes-Gate could serve as a predictive dynamics model within a model-based reinforcement-learning framework, allowing candidate actions to be evaluated according to their predicted downstream glucose consequences before recommendations are delivered. Over repeated observations, the policy could potentially adapt to heterogeneous individual responses and move from population-level prediction toward personalized behavioral guidance.

Another direction is to move from point prediction toward calibrated, risk-aware forecasting. Although MealRes-Gate reduced mean prediction error in extreme-glucose states, absolute errors remained larger in these states than in mid-range glucose windows, precisely where inaccurate forecasts may be most consequential. Deployment-oriented systems should therefore communicate not only a predicted trajectory but also the uncertainty surrounding that prediction. Calibrated prediction intervals, probabilistic forecasts of threshold-crossing events, and abstention mechanisms could distinguish high-confidence predictions from situations in which the system should defer or avoid issuing a recommendation. This consideration becomes even more important when forecasts are embedded within downstream decision-making algorithms, because predictive uncertainty should propagate to the confidence of any suggested action. Future evaluation should therefore extend beyond average forecasting error to examine interval coverage, calibration, event sensitivity and specificity, alert timeliness, decision utility, and the relative consequences of false-positive and false-negative predictions.

## 4 Methods

### 4.1 Study population and participant selection

The FHS and its CGM examination were conducted according to the guidelines of the Declaration of Helsinki and approved by the Boston University Medical Campus Institutional Review Board (protocol number H-41473). All participants provided written consent prior to participation in the study.

The FHS Third Generation, Omni 2, and New Offspring Spouse cohort included 2,718 participants who attended their fourth examination. Exclusions were applied sequentially for study eligibility and device-related reasons, inadequate CGM data quality, and diabetes-related clinical criteria (9). First, 571 participants were excluded because they were ineligible owing to medication use affecting glucose metabolism (*n* = 2), declined CGM wear (*n* = 304), did not return the device (*n* = 203), or returned a device with no transmittable data (*n* = 62).

Second, 128 participants were excluded on the basis of CGM data quality. For this screening step, a CGM day was defined as 6:00 AM to 6:00 AM the following day. We excluded participants with no complete CGM days (*n* = 44), less than 80% of readings within a complete day (*n* = 13), an excessive frequency of glucose values below 70 mg/dL (*n* = 11), or three or fewer full usable days (*n* = 60). Excessively low glucose was defined as values below 70 mg/dL for more than 40% of samples over more than 30% of days, or for more than 50% samples on at least two days.

Third, 267 participants were excluded because diabetes status was missing (*n* = 36), self-reported diabetes was not supported by available biomarkers or medication records (*n* = 5), or diabetes was confirmed by a glycated hemoglobin (HbA1c) ≥ 6.5%, fasting glucose ≥ 126 mg/dL, or glucose-lowering medication use, *n* = 226). The final analytic cohort comprised 1,752 participants.

### 4.2 Forecasting task and evaluation splits

The Dexcom G6 Pro records interstitial glucose every five minutes. Each participant’s CGM series was partitioned into non-overlapping 30-min windows containing six consecutive measurements. The sampling grid was anchored to the participant’s first CGM observation rather than to clock time, so window boundaries were participant-specific.

Each forecasting instance used 12 consecutive windows, representing six hours of glucose history, to predict the next 24 five-minute glucose readings. Predictions were evaluated in four successive 30-minute forecast windows corresponding to 0–30, 30–60, 60–90, and 90–120 min after the forecast origin. Throughout, a *window* refers to a 30-min unit, distinguished by its role as an input or forecast window.

Within each participant, eligible forecasting examples were ordered chronologically and divided into contiguous training (60%), validation (20%), and test (20%) segments without temporal gaps. The same participants therefore contributed to all three segments, and this primary split evaluated temporal generalization within previously observed individuals. Generalization to previously unseen individuals was evaluated separately using five-fold participant-level CV, in which participants were partitioned into five mutually exclusive groups and each fold held one group out, dividing it at the participant level into disjoint validation and test subsets.

All model inputs were restricted to information observed at or before the end of window *t*. In particular, meals recorded at or after the forecast origin were excluded from the input. Labels used only for retrospective subgroup evaluation, including postprandial status, valid-diet-day status, and glucose-zone status, were constructed after prediction and never supplied to the forecasting model.

### 4.3 CGM preprocessing and prediction construction

The Dexcom device has an accuracy floor and ceiling of 40 and 400 mg/dL, respectively, and therefore, reports the strings “Low” and “High” for values outside its measurable range [58]. We recoded the “Low” and “High” as 40 and 400 mg/dL and the recoded values were retained when they occurred in a forecast target. Errors at the sensor limits are therefore evaluated against clamped rather than latent glucose values and may be modestly compressed.

Missing CGM observations were handled before long-horizon feature construction. Missing intervals of at most 90 minutes were filled using up to 10 observed samples on either side of the gap. A cubic smoothing spline was used when at least four context points were present, and linear interpolation was used otherwise. Gaps longer than 90 minutes remained missing. Filled values were only used to calculate historical summary features, including rolling statistics, exponentially weighted summaries, mean amplitude of glycemic excursions (MAGE), and spectral features. They were never used as raw model inputs or as forecast targets.

A forecasting instance was retained only when all raw CGM values in its 12 input windows and all 24 target values were originally observed rather than filled. A 24-hour burn-in period was applied at the start of each participant’s record so that features within a 24-hour lookback could be computed, and forecasts for which any required historical feature remained unavailable were excluded.

Each 30-minute window was represented by a 60-dimensional CGM vector and a 20dimensional dietary vector. The CGM vector contained the six raw glucose readings in the window and 54 engineered features summarizing local dynamics, longer-term glycemic history, and time of day. The dietary vector contained 15 nutrient measures and five meal-context variables. In the full forecasting model, the dietary vector was augmented with the CGM-derived meal probability, giving 21 dietary channels per window. Table 1 summarizes the predictors.

**Table 1:** Predictors constructed for each 30-min window. Dietary predictors describe food reported within the window and recent intake. Short-term CGM predictors are derived from the current window, long-term CGM predictors summarize glucose history extending up to 24 hours before it, and contextual predictors encode time of day. The meal probability is estimated from CGM rather than reported, and is supplied only to the full model.

| Feature class | Category | Variables | Count |
| --- | --- | --- | --- |
| Dietary | Nutrient composition | Energy, net carbohydrate, fiber, total and added sugar, food amount, three fatty-acid classes, whole and refined grains, and percentage of energy from carbohydrate, protein, fat, and alcohol | 15 |
| Dietary | Meal context | Meal indicator, cumulative net carbohydrate over 1 and 2 h, cumulative energy over 2 h, and minutes since the preceding meal | 5 |
| CGM, short-term | Raw window | Six five-minute glucose values in the current window | 6 |
| CGM, short-term | Local distribution | Mean, standard deviation, minimum, maximum, and range | 5 |
| CGM, short-term | Trend and velocity | Linear slope; 5-, 10-, and 25-min changes; lagged-velocity contrast | 5 |
| CGM, short-term | Local excursion | Area below 70 and above 140 mg/dL, and positive and negative area relative to the window mean | 4 |
| CGM, long-term | Rolling summaries | Mean and standard deviation over 1, 2, 3, 6, 12, and 24 h | 12 |

Table 1 (continued)
| Feature class | Category | Variables | Count |
| --- | --- | --- | --- |
| CGM, long-term | Exponentially weighted summaries | Mean and standard deviation at 1-, 2-, 3-, 6-, 12-, and 24-h half-lives | 12 |
| CGM, long-term | Glycemic variability | MAGE over 6, 12, and 24 h | 3 |
| CGM, long-term | Day-over-day reference | 24-h lag and current-minus-24-h change | 2 |
| CGM, long-term | Spectral summaries | Largest PSD ordinate, sum of the PSD ordinates, and their ratio over 6, 12, and 24 h |  |
| CGM, contextual | Circadian phase | Sine and cosine encoding of clock time | 2 |
| CGM-inferred | Meal probability | Probability that a meal occurred in the current window, estimated from the engineered CGM features by the GEE classifier. Derived from CGM rather than from the food record, but supplied to the dietary pathway | 1 |
| **Engineered CGM features** |  |  | **54** |
| **CGM input vector** |  |  | **60** |
| **Reported dietary features** |  |  | **20** |
| **Dietary pathway in the full model, including $\hat{p}^{\text{meal}}$** |  |  | **21** |

#### 4.3.1 Dietary predictors

Food records were aligned to the same 30-minute grid, using data collected from ASA24. Within a window, the 11 absolute-unit food and nutrient variables (food amount, energy, fiber, monounsaturated, polyunsaturated and saturated fat, added sugars, total sugars, whole grains, refined grains, and net carbohydrate) were summed across all food entries. The four macronutrient percentage variables (carbohydrate, protein, fat, and alcohol) are caloric proportions and were combined as energy-weighted averages.

Additional five meal-context variables were included. A binary indicator of whether any caloric food entry was recorded in the window, cumulative net carbohydrate over the preceding one and two hours, cumulative energy over the preceding two hours, and minutes since the most recent recorded meal. The minutes since the most recent meal variable was capped at 360 minutes and scaled to [0,1], taking the value of zero in a window containing a meal and one when no meal occurred earlier in the six-hour lookback.

#### 4.3.2 Short-term CGM predictors

Short-term predictors were computed from the current window. They comprised the six raw glucose values, change in glucose over the preceding 5, 10, and 25 minutes, a lagged velocity contrast, window summary statistics (mean, standard deviation, slope, minimum, maximum, range), and four area-based summaries in mg/dL·min: area above 140 mg/dL, area below 70 mg/dL, and positive and negative areas relative to the window mean.

#### 4.3.3 Long-term CGM predictors

Long-term predictors summarized glycemic history up to 24 hours before the end of the current window. Rolling means and standard deviations were computed over 1, 2, 3, 6, 12, and 24 hours. Exponentially weighted means and standard deviations were computed at the same six half-lives. MAGE was computed over the preceding 6, 12, and 24 hours using the R package iglu (version 4.2.2) [60]. Spectral summaries were obtained by Welch’s method over 6-, 12-, and 24-hour segments [61] and included the largest power spectral density ordinate, the sum of the density ordinates, and their ratio [62]. Two additional contextual predictors captured circadian structure: the glucose value exactly 24 hours earlier and the corresponding day-over-day change. Time of day entered as sine and cosine encodings of the 24-hour clock.

#### 4.3.4 Feature standardization and clipping

Continuous predictors were standardized to zero mean and unit variance using statistics estimated on the training partition only and applied unchanged to validation and test data. In the cross-validation analyses, scaling statistics were re-estimated within each fold’s training partition. Dietary *z*-scores were clipped to [−5, 5], and nutrient channels were set to zero in windows without a recorded food entry, so that absence of food is represented exactly rather than as a scaled zero. The binary meal indicator and the CGM-derived meal probability were left unscaled, both already lying in [0,1].

Forecast targets were left in their original mg/dL units and were never standardized, so that the zone thresholds in the training objective and the reported errors are both expressed directly in clinical units.

### 4.4 Dietary reporting and evaluation strata

#### 4.4.1 Valid diet days

To restrict postprandial analyses to days on which dietary intake was reliably reported, we applied a sex-specific valid-diet-day filter. A calendar day was diet-eligible when total reported energy fell within 650 to 5,700 kcal/day for male participants and 600 to 4,400 kcal/day for female participants [63]. A participant contributed windows only if at least one of their days satisfied this criterion. The flag was propagated to each window by its calendar day and to each forecast horizon by the calendar day of its target. Postprandial and non-postprandial strata are restricted to eligible windows, and the complementary ineligible diet-day stratum reported performance on ineligible windows as a robustness check (Supplementary Figure S2).

Let *a_h_* and *e_h_*= *a_h_* + 30 minutes denote the start and end of forecast window *h*. The window was labeled postprandial when at least one food entry with energy above zero had a reported consumption time in the interval [*a_h_* − 120 minutes*, e_h_*]. This definition includes meals during the two hours preceding the forecast window as well as meals recorded within the window itself.

#### 4.4.2 Glucose-zone strata

Glucose-zone strata form a mutually exclusive and exhaustive partition of each (window, horizon) pair under a low-glucose-priority rule. A forecast window is assigned to the **low** zone if any target value falls below 70 mg/dL, regardless of co-occurring high values. It is assigned to the **high** zone if any target value exceeds 140 mg/dL and the window is not low, and to the **mid** zone otherwise.

### 4.5 CGM-based estimation of meal probability

A window was labeled meal-positive if the participant’s food record contained at least one entry with energy above zero within that window, and meal-negative otherwise.

This is a concurrent detection task: given CGM features from a window and its preceding history, did a meal occur in that same window?

The detector was a generalized estimating equations (GEE) model with a binomial family and logit link, using an exchangeable within-participant working correlation [64]. GEE was used because each participant contributes many temporally correlated windows. It was fitted on the 54 engineered CGM features, using the training partition restricted to valid diet days so that the detector’s training population matches the population on which its output is later consumed. After filtering, this comprised 100,149 windows from 909 of the 1,752 participants, of which 9,953 (9.9%) were meal-positive and 90,196 (90.1%) meal-negative.

Performance was assessed by five-fold participant-level CV, and the ROC-AUC (0.705) is computed from predictions pooled across held-out folds. The coefficients deployed to compute *p̂*^meal^ in the primary temporal-split analyses came from a single final model refit on all valid-diet training windows, so that the cross-validation folds estimated detector generalization while the all-data refit stabilized the deployed coefficients. In the participant-level cross-validation analyses, coefficients were instead re-estimated within each fold’s training participants, so that no held-out participant contributed to the meal probability supplied for their own windows.

Standardized GEE coefficients were summarized descriptively in Supplementary Tables S2 and S1. Positive coefficients indicate higher estimated probability of a concurrent meal, but correlated predictors prevent interpreting individual coefficients as independent feature effects.

### 4.6 Residual-gated Transformer forecaster

The central design principle is that the CGM-based forecast is generated independently of dietary information, with dietary features entering only as an auxiliary correction. This is motivated by the substantial incompleteness of dietary records in populations without diabetes, where meal records are frequently unavailable at inference time.

For a forecast window *t*, the model uses the preceding *T* = 12 windows, *t* − *T* + 1*, . . . , t* as input.

#### CGM forecasting backbone

The backbone is a decoder-based transformer operating exclusively on CGM-derived predictors. The *T* = 12 window feature vectors are projected to dimension *d*, combined with sinusoidal positional encodings, and processed through *L* layers of unidirectional self-attention, implemented with a triangular mask so that each position attends only to itself and preceding positions [65]. The resulting representation **M** ∈ R*^T^ ^×d^* receives no dietary input and therefore yields a complete glucose representation whether or not meals were recorded.

#### Dietary cross-attention pathway

Dietary information enters through a separate cross-attention pathway. The per-window dietary vectors are projected into the same *d*-dimensional space by a two-layer feed-forward network with intermediate dimension *d*_food_, and serve as keys and values, with the CGM representation **M** as queries. Using the CGM state as the query means the dietary signal is retrieved conditional on the current glucose context, so the same recorded meal can yield different corrections depending on where glucose currently sits.

A key-padding mask suppresses windows without a recorded meal. When no window in the lookback contains a recorded meal, the mask is removed rather than applied, which both avoids an undefined attention distribution and allows the CGM-derived meal probability channel to shape the dietary representation when nothing was logged.

The cross-attention output feeds only the dietary correction head, never the CGM head, so the baseline forecast remains dietary-blind. The CGM head produces a 24-value baseline forecast **ŷ**cgm, and the dietary head produces an additive correction Δ**ŷ**^food^ of the same dimension. The final layer of the dietary head is initialized to zero, so the model begins as a purely CGM-based forecaster and develops a dietary pathway only where supported by the data.

#### Meal-aware residual gating

A gating network scales the dietary correction separately at each of the four forecast windows using two dietary features from the most recent input window, window *t*: minutes since the most recent recorded meal and cumulative net carbohydrate over the preceding two hours. The gate is

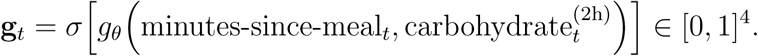

where *g_θ_* is a two-layer feed-forward network. Its four sigmoid gate biases were initialized to −3, giving initial values of approximately 0.047. Combined with the dietary head’s zero-initialized output layer, which makes the dietary correction exactly zero at initialization, the near-closed gate initially keeps the dietary corrections small until the model learns to increase the gate values.

Let 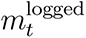 indicate that at least one meal was recorded in windows *t* − *T* + 1*, . . . , t*, that is, anywhere in the six-hour lookback, and let *p̂*^meal^ be the detector’s probability for the current window. The effective meal-presence signal is

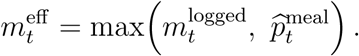

A recorded meal therefore opens the pathway fully and is never attenuated by a low inferred probability, while the inferred probability can still open it partially when nothing was recorded.

The final forecast is

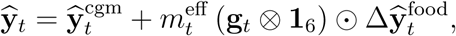

Thus, each of the four gate values scales the six successive five-minute dietary corrections in its corresponding forecast window.

### 4.7 Training and hyperparameter selection

#### Training objective

The objective combined a zone- and horizon-weighted smooth *L*_1_ prediction loss with a velocity-consistency loss [66]. For a mini-batch of size *B*, the prediction loss was

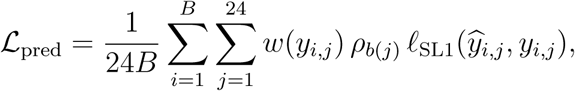

where *b*(*j*) = ⌈*j/*6⌉ identifies the forecast window, *ρ* = (1.0, 0.8, 0.6, 0.4) gives the four horizon weights, and *w*(*y*) equals 2.0 for *y <* 70 mg/dL, 1.5 for *y >* 180 mg/dL, and 1.0 otherwise. The smooth *L*_1_ transition parameter was 1.0.

The velocity-consistency loss compared successive changes in the predicted and observed trajectories:

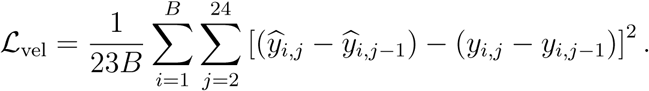

The total objective was L = L_pred_ + *λ*_vel_L_vel_, with *λ*_vel_ = 0.1. The velocity term was not multiplied by the zone or horizon weights and therefore acted uniformly across the trajectory.

The *>* 180 mg/dL threshold in the objective is intentionally different from the *>* 140 mg/dL threshold defining the high-glucose evaluation stratum. The former marks clinically pronounced hyperglycemia during optimization, whereas the latter gives a more sensitive evaluation partition for a cohort without diabetes.

#### Optimization and selection

Hyperparameters were selected by Bayesian optimization with Optuna [67] using a tree-structured Parzen estimator sampler, maximizing the validation coefficient of determination (*R*^2^). Early stopping and learning-rate schedule used the same validation criterion. Models were optimized with AdamW [68]. The selected configuration was refit for at most 200 epochs with early stopping. Search ranges and selected values are given in Supplementary Tables S8 and S9. Baseline hyperparameters were selected separately on the same validation partition using architecture-specific search spaces and search budgets.

For every model, the selected configuration was held fixed across random seeds, ablation variants, and cross-validation folds. No hyperparameter was retuned per seed or per fold. All multi-seed results use five random seeds.

### 4.8 Comparison models

Three neural forecasting architectures were compared. The first was an LSTM, representing recurrent approaches common in glucose forecasting [69, 37]. The second was a standard Transformer applying self-attention over the same six-hour CGM history [53, 70]. The third was the GluFormer architecture [41], introduced as a generative foundation model pretrained on more than 10 million glucose measurements; here its architecture was trained from scratch on the study data because the pretrained weights are not publicly available.

All baselines used the same partitions, forecast targets, and evaluation as the proposed model, received only raw CGM input without engineered or dietary predictors, and were trained under a mean-squared-error objective. This is therefore a framework-level comparison under matched study data and evaluation conditions, rather than an architecture-only comparison or a reproduction of an externally pretrained model.

### 4.9 Forecast evaluation

#### 4.9.1 Glucose prediction error

For each 30-min forecast window, window-averaged mean absolute error (MAE) was defined as the mean of the six pointwise absolute errors between predicted and observed five-minute glucose values. Overall and stratum-specific performance was obtained by aggregating these window-level values; for participant-level analyses, MAE was first averaged across all eligible windows contributed by each participant.

Valid-diet-day, postprandial, and glucose-zone labels were assigned only after forecasts were generated. The postprandial label may therefore reference meals occurring during the forecast interval, and glucose-zone labels use observed target values, because these variables define retrospective evaluation strata only and never entered model fitting or selection.

#### 4.9.2 Postprandial trajectory metrics

For cumulative horizons *H* ∈ {30, 60, 90, 120} minutes, the magnitude, timing, and duration of the postprandial trajectory were summarized [5, 19]. Let *z*(*u*) denote an observed or predicted trajectory over 0 *< u* ≤ *H*, and let *b* be the final observed CGM value before the forecast origin. These metrics were evaluated only on forecasts for which a meal was recorded in the 30-minute input window immediately preceding the forecast origin.

Peak glucose was max_0_*_<u≤H_ z*(*u*) and time to peak the elapsed time to the maximizing observation, taking the earliest observation when the maximum was attained more than once. The pAUC was

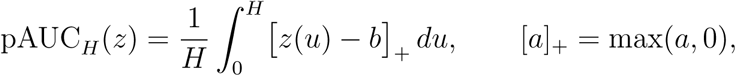

evaluated by the trapezoidal rule over the five-min observations. Dividing by *H* expresses pAUC in mg/dL as a time-averaged excursion above baseline, making values comparable across cumulative horizons. Negative deviations are truncated at zero to isolate the excursion component [71, 72]. Time in range was the proportion of trajectory values satisfying 70 ≤ *z*(*u*) ≤ 140 mg/dL [73]. Predicted and observed values of each metric were compared by absolute error.

### 4.10 Statistical analysis

Statistical tests were performed at the participant level. statistical significance was assessed at *α* = 0.05 after multiplicity adjustment where applicable.

#### 4.10.1 Statistical testing used in Section 4.2 and 2.7

MealRes-Gate was compared separately with the transformer, LSTM, and GluFormer models within each evaluation stratum and forecast horizon. Within each training seed, MAE was first averaged over the same eligible test windows for each participant, and the participant-level paired difference was calculated as comparison-model minus MealRes-Gate MAE. For participant-level cross-validation, only predictions from the fold in which a participant was held out for testing were used, and the nonoverlapping test subsets were combined within each seed.

Participant-level paired differences were tested against zero within each seed using twosided Wilcoxon signed-rank tests [74]. For each comparison, a 5-of-5 partial-conjunction *p*-value was defined as the maximum of the five unadjusted seed-specific *p*-values, thereby requiring evidence in all five seeds [75, 76]. Each evaluation design consisted of 72 comparisons (three comparison models by six strata by four forecast horizons). The resulting conjunction *p*-values were adjusted using the Benjamini–Hochberg procedure [77], separately for the temporal-split and participant-level cross-validation evaluations.

#### 4.10.2 Statistical testing used in Section 2.4

Subgroup analyses were performed using results from the single training run show in Figure 4. At each forecast window, participant-level MAE was obtained by averaging all eligible window-level values for each participant. These distributions are right-skewed, so rank-based tests were used.

Sex and prediabetes status were analyzed as two-group variables using two-sided Mann– Whitney *U* tests [78]. Age, body mass index, glucose coefficient of variation, and the percentage of CGM readings above 140 mg/dL were divided into equal-frequency tertiles and evaluated using Kruskal–Wallis tests [79]. The resulting 24 primary *p*-values, corresponding to six subgroup dimensions across four forecasting windows, were adjusted using the Benjamini–Hochberg procedure [77]. Statistical significance in Figure 4 was determined from these adjusted primary *p*-values.

As a post hoc assessment of where differences occurred among tertiles, pairwise contrasts were evaluated using two-sided Mann-Whitney *U* tests with pairwise *p*-values adjusted using the Benjamini–Hochberg procedure separately within each subgroup dimension and forecast window.

## Data availability

The Framingham Heart Study (FHS) data analyzed in this study will be uploaded to the following repositories during the next FHS data transfer: BioLINCC (https://biolincc.nhlbi.nih.gov/home/), dbGaP (https://www.ncbi.nlm.nih.gov/projects/ gap/cgi-bin/study.cgi?study_id=phs000007.v33.p14), and BioData Catalyst (https://biodatacatalyst.nhlbi.nih.gov/). FHS is also available upon request through the Research Application on the FHS website (https://www.framinghamheartstudy.org/).

## Code availability

Implementation of MealRes-Gate model architecture is available at https://github.com/jungscode/MealRes-Gate

## Data Availability

The Framingham Heart Study (FHS) data analyzed in this study will be uploaded to the following repositories during the next FHS data transfer: BioLINCC, dbGaP, and BioData Catalyst. FHS data are also available upon request through the Research Application on the Framingham Heart Study website.

https://biolincc.nhlbi.nih.gov/home/

https://www.ncbi.nlm.nih.gov/projects/gap/cgi-bin/study.cgi?study_id=phs000007.v33.p14

https://biodatacatalyst.nhlbi.nih.gov/

https://www.framinghamheartstudy.org/

## Acknowledgements

This work was supported by the Framingham Heart Study’s National Heart, Lung, and Blood Institute contracts (N01-HC25195, HHSN268201500001I, and 75N92019D00031), and the National Institute of Diabetes and Digestive and Kidney Diseases (R01DK129305). Additional support was provided by the National Institute of General Medical Sciences (NIGMS) Interdisciplinary Training Program for Biostatisticians (T32 GM140972).

Dexcom provided continuous glucose monitors at a discounted rate.

## Author contributions

HC and JL conceived the study. NLS contributed to data acquisition. JL and SY performed the data analysis. HC and JL drafted the manuscript. All authors contributed to interpreting the results and revising the manuscript. All authors reviewed and approved the final manuscript.

## Competing interests

NLS serves on a Scientific Advisory Board for Abbott and received travel support and honorarium for her services. Other authors declare no competing interests.

## Notes

### Author Declarations

Boston University Medical Campus Institutional Review Board gave ethical approval for this work (protocol number H-41473).

### Summary of Updates

This version includes updates to the title, abstract, methods, results, figures, and discussion. We also clarified the MealRes-Gate model and evaluation procedures, updated author information and disclosures, and revised the funding and data and code availability statements.

